## Supplemental Figures and Tables for "*ADRA2A* and *IRX1* are putative risk genes for Raynaud’s phenomenon"

[1]Computational Medicine, Berlin Institute of Health at Charité – Universitätsmedizin Berlin, Berlin, Germany
[2]Preventive Neurology Unit, Wolfson Institute of Population Health, Queen Mary University of London, London, UK
[3]Institute of Health Informatics, University College London, London, UK
[4]Health Data Research UK, London, UK
[5]British Heart Foundation Data Science Centre, London, UK
[6]National Institute of Health Research University College London Hospitals Biomedical Research Centre
[7]Department of Pharmacology and Therapeutics, The Wolfson Centre for Personalised Medicine, University Liverpool, Liverpool, UK, Liverpool, UK
[8]Vanderbilt Genetics Institute, Vanderbilt University Medical Center, Nashville, TN 37203, USA
[9]William Harvey Research Institute, Queen Mary University of London, London, UK
[10]MRC Epidemiology Unit, University of Cambridge, Cambridge, UK
[11]Precision Healthcare Institute, Queen Mary University of London, London, UK
*these authors jointly supervised the work

**Correspondence**

Prof Claudia Langenberg
Dr Maik Pietzner

### Supplementary Figures

#### Supplementary Figure 1 – Inclusion criteria and primary RP definition

**Flowchart for the criteria for inclusion** into the study and criteria for definition as primary Raynaud's phenomenon.

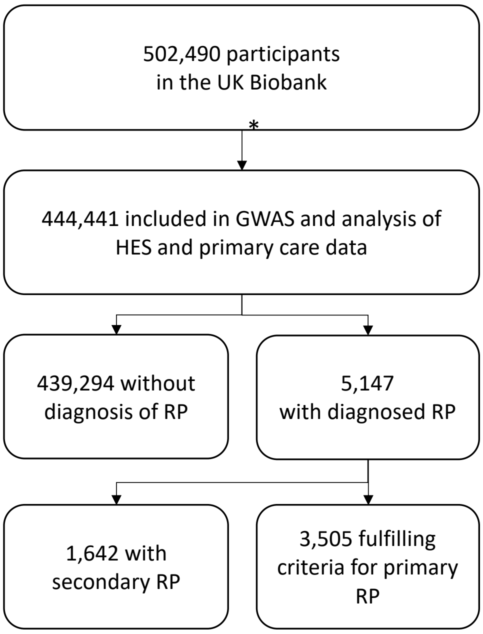

Criteria for primary Raynaud Phenomenon:

Prior and later to the diagnosis of RP the following diseases have never been diagnosed:

- Arteriosclerosis
- Dermatomyositis
- Polymyositis
- Systemic sclerosis
- Systemic Lupus Erythematosus
- Sjoegren Syndrome
- Diseases of connective tissue
- Cryoglobulinemia
- Autoimmune haemolytic anaemias
- Carpal tunnel syndrome
- Frostbite
- Hypothyroidism
- Medication with beta blockers

RP = Raynaud phenomenon, HES = hospital episode statistics
*Excluded due to missing genetic data, genetic data did not pass quality control, or withdrawn consent

### Supplementary Figure2 – Regional association plot

Regional association plot for variants reported in table 1 for Raynaud's Phenomenon (RP). Each plot displays p-values from logistic regression models associating single nucleotide variants in a ±500kb window around the lead signal with the risk of RP. The colour gradient indicates the linkage disequilibrium ($r^2$) with the lead signal (annotated) at the respective locus.

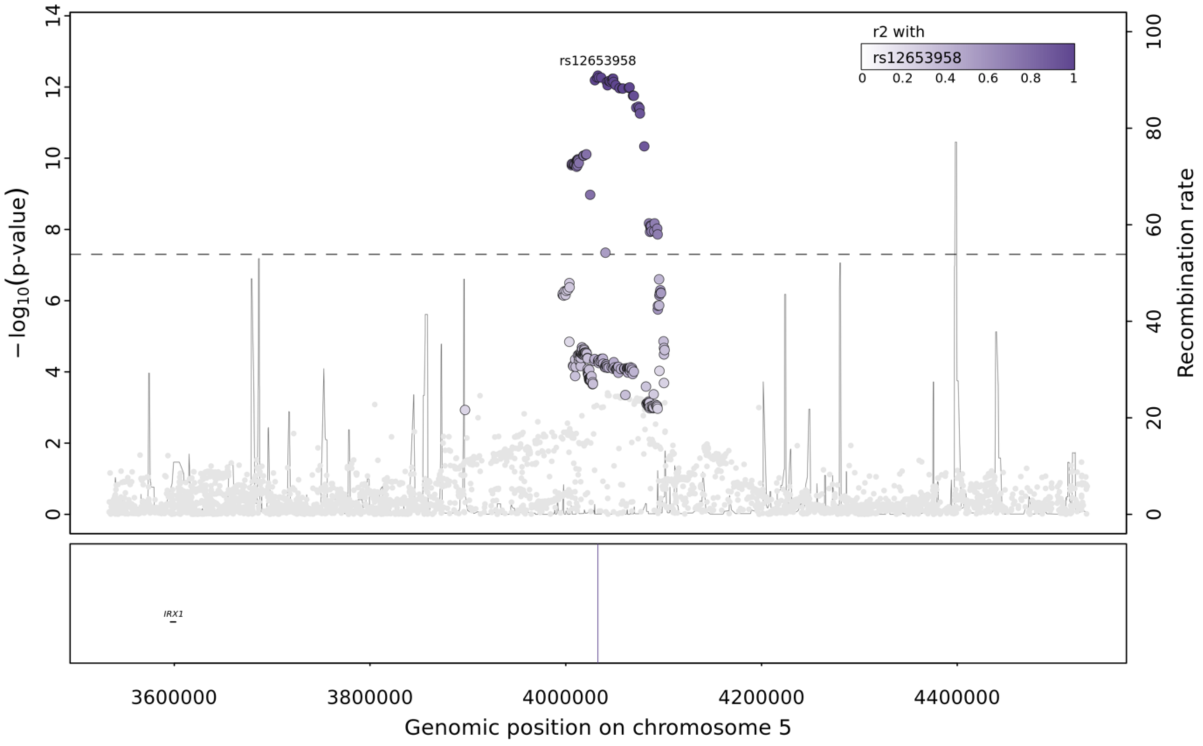

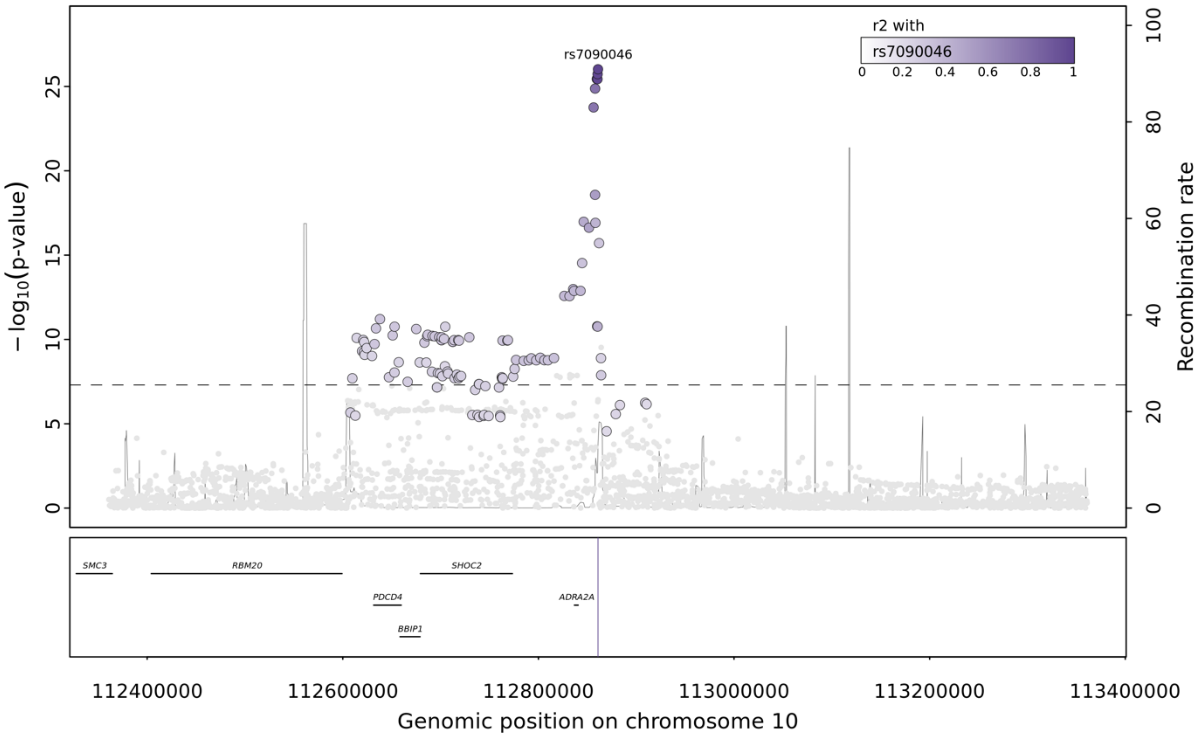

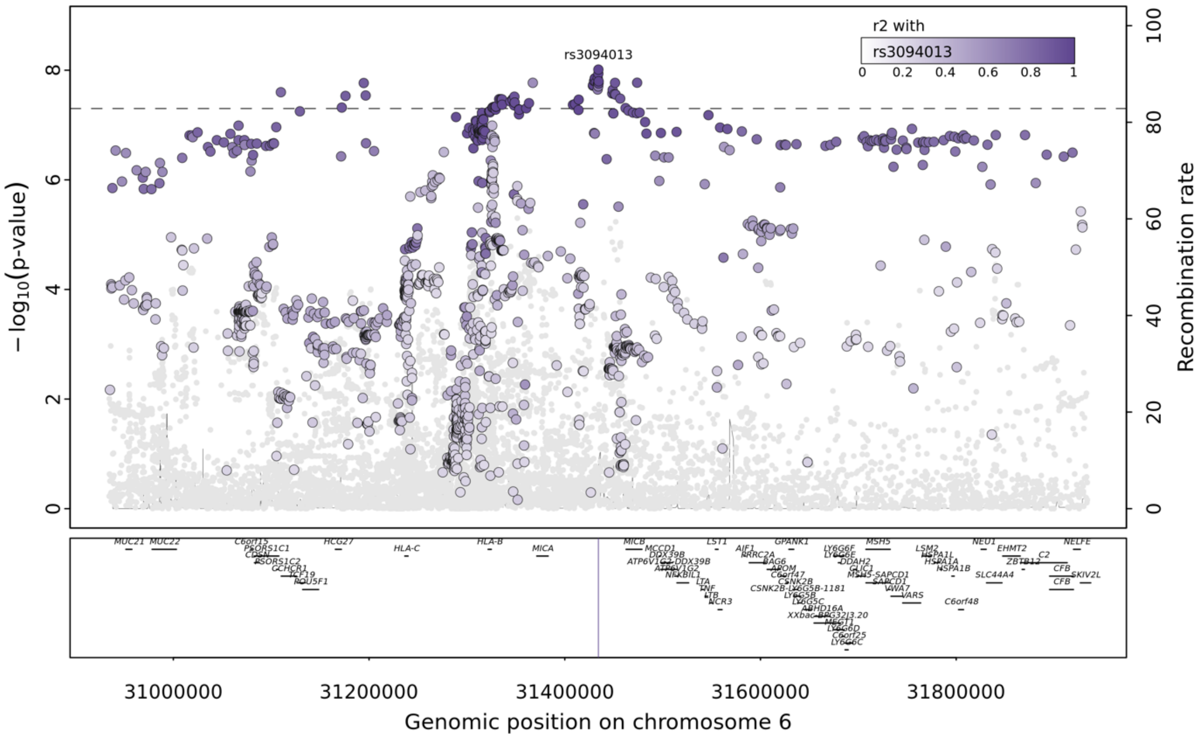

### Supplementary Figure 3 – Q-Q plot

Q-Q plot

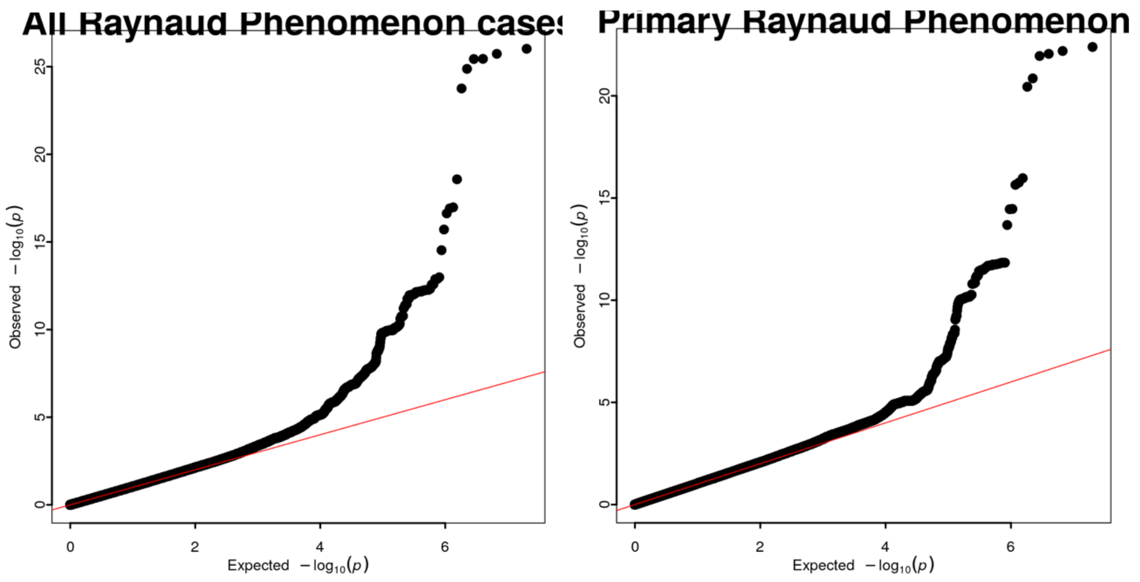

**Supplementary Figure 3** QQ-plot of the genome-wide association results for all RP cases (left) and restricting to primary RP cases (right).

### Supplementary Figure 4 – Beta-correlation plot

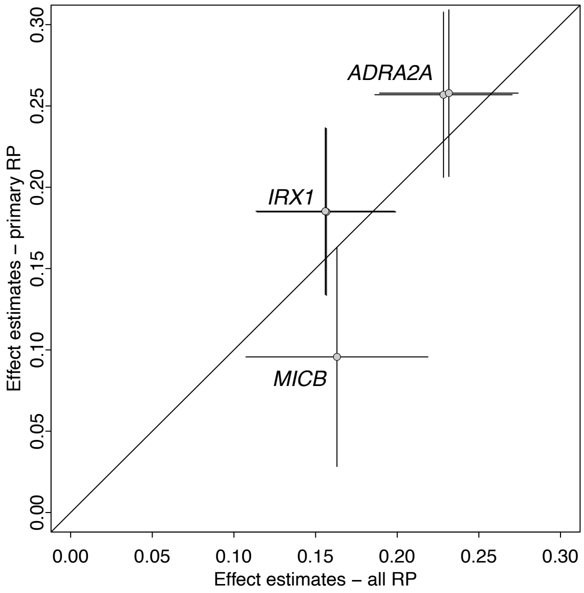

**Supplementary Figure 4** Comparison of effect estimates for three genetic loci, five distinct variants, significantly associated with RP risk among all RP cases (x-axis) and when restricting to primary RP cases only (y-axis).

### Supplementary Figure 5 – Genetic correlation plot

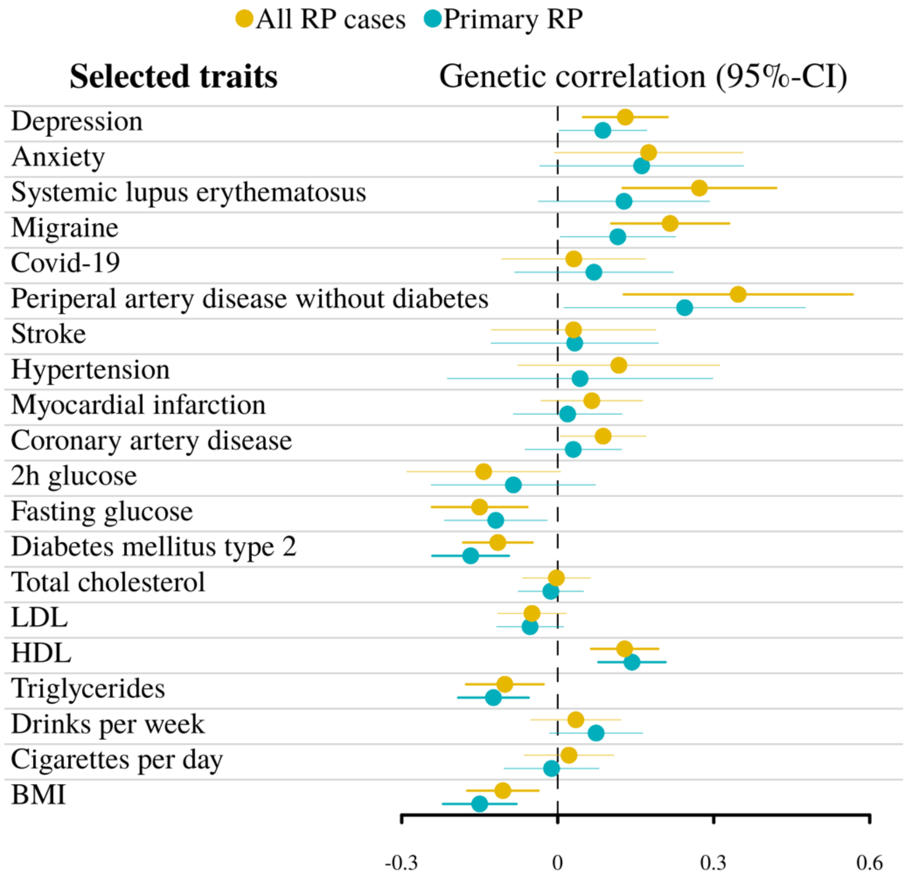

**Supplementary Figure 5 Forest plot summarizing genetic correlation analyses between RP and selected traits** Results for selected traits as listed in Supplementary Data 2. Dots indicate point estimates for genetic correlations and lines indicate 95%-confidence intervals.

### Supplementary Tables

#### Supplementary Table 1 – Characteristics of cases and controls

**Supplementary Tab. 1:** Characteristics of case and control population  Sample size, demographic factors, and comorbidities of Raynaud's phenomenon (RP) cases and controls. P values were obtained using Chi-Square test for categorical variables and ANOVA for continuous variables. All p-values are two-sided.

| *Variable (cases/controls (in case participants did not declare an answer))* | *Participants with Raynaud's phenomenon* | *Controls* | *p-value* |
|---|---|---|---|
| *n* | 5,147 | 439,294 | |
| *Age in years (mean (SD))* | 57.84 (7.9) | 56.80 (8.0) | $5.9 \times 10^{-7}$ |
| *Sex (%) [Male]* | 1,639 (31.8) | 201,704(45.9) | $<2.2 \times 10^{-16}$ |
| *BMI in m2/kg (mean (SD))* | 25.86 (4.6) | 27.42(4.8) | $<2.2 \times 10^{-16}$ |
| *Smoking (%) (9048/433817)* | | | $4.3 \times 10^{-7}$ |
| *Never* | 2,493 (50.6) | 236,881 (54.1) | |
| *Ever* | 2,535 (49.4) | 200,856 (45.9) | |
| *Alcohol (%)* | | | $6.5 \times 10^{-4}$ |
| *Never* | 205 (4.0) | 13,789(3.1) | |
| *Ever* | 4,937 (96.0) | 425,114 (96.9) | |
| *Diastolic blood pressure in mmHg (mean (SD))* | 80.49 (10.6) | 82.22 (10.7) | $<2.2 \times 10^{-16}$ |
| *Systolic blood pressure in mmHg (mean (SD))* | 138.91 (20.2) | 140.03 (19.6) | $1.3 \times 10^{-4}$ |
| *Town Deprivation Index (mean (SD))* | -1.42 (3.0) | -1.49 (3.0) | 0.131 |
| *Household income in Pound (%)* | | | $<2.2 \times 10^{-16}$ |
| *Less than 30,999* | 2,527 (49.3) | 180,752 (41.3) | |
| *31,000 to 100,000* | 1,658 (32.3) | 177,331 (40.5) | |
| *Greater than 100,000* | 116 (2.3) | 20,216 (4.6) | |
| *Not declared* | 827 (16.1) | 59,498 (13.6) | |
| *Migraine (%)* | 519 (10.1) | 19,287 (4.4) | $<2.2 \times 10^{-16}$ |
| *Hypertension (%)* | 2,190 (42.5) | 144,872 (33.0) | $<2.2 \times 10^{-16}$ |
| *Type 1 Diabetes mellitus (%)* | 60 (1.2) | 4,058 (0.9) | $8.4 \times 10^{-2}$ |
| *Type 2 Diabetes mellitus (%)* | 381 (7.4) | 36,770 (8.4) | 0.014 |
| *Other chronic ischemic heart disease (%)* | 601 (11.7) | 29,892 (6.8) | $<2.2 \times 10^{-16}$ |
| *Systemic Lupus Erythematosus (%)* | 131 (2.5) | 528 (0.1) | $<2.2 \times 10^{-16}$ |
| *Systemic Sclerosis (%)* | 229 (4.4) | 148 (0.0) | $<2.2 \times 10^{-16}$ |

### Supplementary Table 2 – OR for sex and sex*SNP interaction

**Supplementary Tab. 2**: OR for variants for men and women separatly as well as p-value for sex*SNP interaction term. Statistics were derived using logistic regression models.

| SNP | Alleles | Frequency | GWAS | OR women | CI lower women | CI upper women | p-value women | OR men | CI lower men | CI upper men | p-value men | p-value interaction |
|---|---|---|---|---|---|---|---|---|---|---|---|---|
| ***rs7090046*** | A/G | 0.31 | all | 1.28 | 1.21 | 1.36 | 1.01e-18 | 1.20 | 1.11 | 1.31 | 7.97e-06 | 0.20 |
| ***rs12653958*** | A/G | 0.30 | all | 1.14 | 1.07 | 1.20 | 8.58e-06 | 1.20 | 1.11 | 1.30 | 1.23e-05 | 0.28 |
| ***rs3094013*** | G/A | 0.14 | all | 1.15 | 1.07 | 1.24 | 1.24e_04 | 1.23 | 1.11 | 1.37 | 6.48e-05 | 0.31 |
| ***rs1343449*** | A/G | 0.32 | primary | 1.30 | 1.22 | 1.39 | 3.64e-14 | 1.19 | 1.09 | 1.30 | 1.71e-04 | 0.13 |
| ***rs72731435*** | T/C | 0.30 | primary | 1.17 | 1.09 | 1.26 | 9.39e-06 | 1.23 | 1.12 | 1.35 | 9.88e-06 | 0.41 |

OR = odds ratio; CI = confidence interval; GWAS = type of outcome, all – all RP cases, primary – excluding secondary RP cases

### Supplementary Table 3 – Effect estimates for all and primary RP cases

**Supplementary Tab. 3:** Comparison of effect estimates for genetic variants associated with RP between all and primary cases. Statistics were derived from logistic regression models.

| Variant | | | | | GWAS of all RP (N = 444,441, 5147 cases and 435,357 controls) | | | GWAS of Primary RP(N = 441,542, 3505 cases and 435,357 controls) | | |
|---|---|---|---|---|---|---|---|---|---|---|
| **SNP** | **Chromosome number** | **Position** | **Alleles** | **Frequency** | **OR** | **CI** | **P value** | **OR** | **CI** | **P value** |
| rs12653958 | 5 | 4032849 | A/G | 0.30 | 1.17 | (1.12;1.22) | 4.76E-13 | 1.20 | (1.14;1.27) | 1.69E-12 |
| rs72731435 | 5 | 4048526 | T/C | 0.30 | 1.17 | (1.12;1.22) | 5.85E-13 | 1.20 | (1.14;1.27) | 1.46E-12 |
| rs3094013 | 6 | 31434366 | G/A | 0.14 | 1.18 | (1.11;1.24) | 9.73E-09 | 1.10 | (1.03;1.18) | 5.40E-03 |
| rs1343449 | 10 | 112860526 | A/G | 0.32 | 1.26 | (1.20;1.31) | 1.85E-26 | 1.29 | (1.23;1.36) | 4.04E-23 |
| rs7090046 | 10 | 112860930 | G/A | 0.31 | 1.26 | (1.21;1.32) | 9.58E-27 | 1.29 | (1.23;1.36) | 6.39E-23 |

### Supplementary Table 4 – Latent causal variable analysis

Results from latent causal variable analysis for phecodes with evidence of significant genetic correlations (rg)

| ***T1*** | ***Phecodes*** | ***rg*** | ***SE(rg)*** | ***p.fdr(rg)*** | ***GCP*** | ***SE(GCP)*** | ***p.fdr(GCP)*** |
|---|---|---|---|---|---|---|---|
| *All RP cases* | Osteoporosis NOS | 0.40 | 0.08 | 1.36E-04 | 0.72 | 0.19 | 1.58E-03 |
| *Primary RP* | Osteoporosis NOS | 0.35 | 0.09 | 7.93E-03 | 0.66 | 0.22 | 1.17E-02 |
