## Supplementary material for "*ADRA2A* and *IRX1* are putative risk genes for Raynaud’s phenomenon": Legends for Supplementary Data Sets

**Supplementary Data Legends**

File Name: Supplementary Data 1
Description: Results from cis-eQTL mapping for regional sentinels for Raynaud's syndrome; Statistics for RP were derived from logistic regression models and statistics for gene expression derived from linear regression models.

File Name: Supplementary Data 2
Description: Sources of publicly available GWAS

File Name: Supplementary Data 3
Description: Results from genetic correlation analysis using selected traits

File Name: Supplementary Data 4
Description: Results from genetic correlation analysis across 185 phecodes with two-sided tests of significance.

File Name: Supplementary Data 5
Description: Summary of genetically prioritized drug targets and drugs in trials for Raynaud's phenomenon

File Name: Supplementary Data 6
Description: Electronic health records codes for identification of used conditions and diseases
